# Improving Medicare Fraud Detection Accuracy in Deep Learning by Exploring Feature Selection and Data Sampling Techniques

**DOI:** 10.64898/2026.03.18.26348763

**Authors:** Fahad Ahammed, Bayan Al Barakati, Oge Marques

## Abstract

Fraud in the health landscape is an aggravating issue, with far-reaching consequences burdening the financial stability of the health industry and threatening the quality of medical care. It results from vulnerabilities within the current healthcare framework that are exploited by the fraudsters in their favor. In spite of many developed models that aim to detect fraudulent patterns in insurance claims, the accuracy of such models frequently suffers as a result of the imbalance issue of the Medicare dataset and irrelevant features. This study ventures to improve detection performance and accuracy by employing a deep learning model along with data sampling and feature selection techniques. Comparative analysis among different combinations is conducted to determine their efficacy to enhance the accuracy of the fraud detection model. Hence, the suggested model clearly demonstrates that a combination of myriad data sampling and feature selection techniques is helping to improve accuracy and performance. The accuracy was thus 95.4%, with negligible evidence of overfitting detected using both Chisquare and Synthetic Minority Over-sampling (SMOTE) techniques. Ultimately, the study findings underscore the significance of employing combined techniques instead of using only the baseline deep learning model for better performance in detecting Medicare insurance fraud.

## 1 Introduction

Healthcare fraud insurance has been a long-standing issue. Primarily, in this age of digitalization, fraudulent activities become more sophisticated, and the traditional detection and prevention mechanisms prove their insufficiency. Across the globe, a massive amount of money is lost to fraud and misrepresentation, which overhead the insurance companies with billions of dollars. In 2021, an estimated loss of over 1.4 billion US dollars as a result of fraud was reported by the National Enforcement Action on Health Care Fraud [1]. Healthcare insurance fraud not only results in mere financial losses, but also puts patient safety and privacy at risk and compromises the credibility of the healthcare system as a whole. Based on Health Care Fraud National Enforcement Action of 2021, a concerning number of more than 12 million opioid tablets were reportedly prescribed, reflecting the scope of the problem being investigated [1]. Medical insurance fraud manifests in variant practices. These include the practice of up-coding services and billing for non-existent treatments or charges for unnecessary services. Moreover, illegal practices include stealing patient identity to acquire benefits illegally, misrepresenting services or procedures, forging or altering the prescriptions, and lastly, over-billing for medical equipment that has never been bought. From a theoretical aspect, study [2] illustrates that fraudulent insurance claims usually have many visits and increased medical bills and expenses compared to genuine claims.

Due to the marked impact of fraud in the medical sector on patient health, trust, system operation, and economics, a considerable number of studies have proposed distinct solutions to detect fraudulent insurance claims. According to [3], classical detection methods like Logistic Regression, Support vector machines(SVMs), and Random Forests work efficiently with balanced data; however, with imbalanced data, this is not the case. Therefore, to overcome the imbalanced data problem, a deep boosting decision tree (DBDT) is a novel model proposed by [3]. The proposed model integrates the neural networks into gradient boosting to enhance its capacity for representation learning while preserving interpretability. Furthermore, using an advanced emerging technology such as blockchain is a hot research area thanks to its decentralized network. The researchers in [4] proposed blockchain and AI-based secure systems to detect illegal insurance fraud. The proposed system consists of four layers, which are the user layer, data generation layer, data analytical layer, and blockchain layer. In the same context, the study [5] combined blockchain with machine learning techniques like XGBoost, Random Forest, Logistic Regression, and Decision Tree to identify the fraud insurance activities. Further, study [6] considered the spatial and temporal aspects of the data, which the orthodox detection methods may not take into account. To do so, the authors proposed a fraud detection approach by employing a spatiotemporal constraint graph neural network (StGNN). The finding indicates that the proposed approach outperformed the traditional detection solutions in terms of accuracy and F1 score.

The main focus of this paper is improving the accuracy of detecting fraudulent patterns in Medicare claims by utilizing a deep learning model alongside feature selection and data sampling techniques. Both techniques were utilized for the following reasons. First, by removing the redundant features, the model can detect insurance claims fraud more accurately. Secondly, by employing the feature selection technique, the model becomes less complex and less overfitting. Additionally, data sampling techniques, such as over-sampling and under-sampling, are able to address the imbalance data issue. Consequently, the proposed model proves that employing a combined model comprising data sampling and feature selection techniques helps to enhance the accuracy and performance explicitly. Thus, the recorded accuracy is 95.4%, with negligible evidence of overfitting detected resulting from utilizing both Chi-square and SMOTE techniques.

This paper is organized as follows: First, Section 2 discusses the literature review related to detecting Medicare fraud. Afterwards, the research methodology is introduced in Section 3. Then, in Section 4, a comprehensive analysis of the obtained results is conducted. Finally, the paper ‘s conclusion and future works are discussed in Section 5.

## 2 Literature Review

In the health insurance sector, the risks of fraudulent claims look threatening, posing a significant challenge to the healthcare system and insurance companies. Thus, the fraudulent activities not only threaten financial resources but also raise ethical and patient safety concerns. Consequently, implementing effective fraud detection mechanisms is crucial to maintain the integrity of the healthcare system and guarantee patient care and well-being.

Initially, deep learning has been utilized alongside blockchain technology in study [7]. The blockchain has been used as secure, tamper-proof, and resistant storage for the medical record. Moreover, the BERT-LE model was developed based on deep learning techniques. This model was designed to evaluate the disease diagnosis validity by anticipating the accuracy of the diagnostic code (ICD disease code). Hence, the researchers used BERT (Bidirectional Encoder Representations from Transformers)a pre-trained natural language processing (NLP)- and also the Label Embedding (LE) layer to encode the input text and anticipate the likelihood of diagnostic codes. This study achieved high effectiveness in identifying and detecting suspicious insurance claims with 87%, which helped significantly reduce the workload of the insurance auditors. To predict medical insurance fraud, research [8] discusses employing the Extreme Gradient Boosting decision tree (XGB) algorithm in order to improve the accuracy of the decision tree classifier. The XGB algorithm was used to train the model and also to detect the potentially fraudulent transactions on the dataset. The XGB tree was implemented by employing a gradient boosting framework, which integrates the predictions of multiple learners, such as decision trees, to create a substantial and accurate predictive model. To evaluate the model, the Random Forest model was used as a baseline model, and the result was compared in terms of accuracy and recall. Therefore, the XGB tree classifier achieved high performance with a random under-sampling method with tuned parameters, except for noise in the data, which worsens the XGB performance and leads to overfitting.

Furthermore, the constrained availability of a labeled dataset in the medical field has long been a challenge. The researchers in [9] and [10] applied innovative solutions to overcome the scarcity of labeled data. The unsupervised multivariate analysis model was proposed by [9]. They used Weighted multi-tree (WMT) to capture the similarities and patterns by analyzing multivariate categorical data. Moreover, to identify anomalies and catch fraudulent claims, Density-Based Clustering (DBC) techniques were employed.

Likewise, the researchers in paper [10] introduced a machine learning-based mechanism for capturing fraud in medical insurance. Therefore, they combined the data sampling method and semi-supervised approach beside the Bagging of Decision Tree (BoDT) regression algorithm. The data sampling addressed the imbalance dataset issue and enhanced data representation by selecting nominal attributes that correlate most with the target features and creating a new composite attribute. Additionally, the semi-supervised approach was utilized to attain historical knowledge about the claims, either fraud or legitimate, to anticipate the deduction rate and detect defrauding claims. Ultimately, the evaluation result showed that BoDT achieved a lower Mean Absolute Error (MAE) compared to the Random Forest model.

Additionally, different data sampling techniques were evaluated on an imbalanced dataset regarding misrepresenting activities in medical insurance claims by [11]. These techniques are Random Under-Sampling (RUS), Random Over-Sampling (ROS), and Synthetic Minority Over-sampling Technique (SMOTE). The comparison of these techniques was achieved over two learners, Random Forest (RF) and Decision Tree. The researchers found that a class ratio 50:50 is not helpful for vast datasets such as Medicare. However, they suggested that a larger ratio of fraud classes (10:90) is scoring better performance. Also, they concluded that the Under-sampling technique is the most effective on the imbalanced dataset. Further, the Random Forest model is more beneficial than the decision tree for detecting fraud claims in medical insurance. For better performance of clustering algorithms and reducing the hospital dataset dimensions, the feature selection algorithm was utilized by [12]. The proposed unsupervised clustering model was designed for healthcare fraud and anomaly detection. It involved two steps: the feature selection and clustering. The feature selection purpose is to opt a subset of the most relevant and appearing features from the dataset. The clustering step groups the data into sets based on the selected features. In addition, feature subset selection based on a squirrel optimization algorithm was employed along with nearest neighbor classification to identify the fraudulent behavior in health insurance activities [13]. Utilizing feature selection led to optimal classification performance regardless of the irrelevant features and rate of classification errors. The result indicated that the proposed model achieved a higher F-measure rate than other classification models that do not employ the feature selection method.

While the discussed studies have made significant progress using traditional machine learning or isolated sampling methods, a gap remains in effectively combining feature selection with synthetic oversampling specifically for deep learning architectures. This work distinguishes itself by integrating Chi-squared feature selection and SMOTE into a unified deep learning pipeline. This combined approach is specifically designed to handle high-dimensional, imbalanced Medicare data, providing a more robust and sensitive detection mechanism than conventional baseline models.

## 3 METHODOLOGY

### 3.1 Dataset

The Dataset used in this paper serves as a key component for the experimental research carried out in this study as it provides the necessary insights and assists in drawing substantial conclusions. The dataset was initially obtained from Kaggle by Rohit Anand Gupta [14] and processed further. It has 5012 unique providers, around 203,000 beneficiaries, and 56 independent features, along with 558,212 claims or sample data. The data is divided into four subsets: Provider details, Beneficiary details, Outpatient details, and Inpatient details. This specific dataset was selected because it provides a comprehensive, multi-dimensional benchmark for Medicare fraud research. Its complexity, involving over half a million merged claims across multiple categories, allows for a rigorous evaluation of how deep learning models handle high-volume, real-world healthcare billing irregularities. As the dataset came into four parts, all the data was merged into a single DataFrame, “Allpatientdata”, by performing outer joins on multiple columns to consolidate information from different sources.

In the course of data preprocessing, a number of new features have been derived from existing data within the Medicare dataset, like patient age, gender, admission data, chronic conditions of the patient, physician, and claim duration. Through the process of grouping and aggregating the dataset, it crystallizes essential statistics such as the mean and standard deviation of particular features. This consolidation is achieved by combining data points based on both the “Provider” and “PotentialFraud” columns, which significantly reduces the dataset size, making it more manageable for training the model. The following bar chart Figure [1] illustrates the distribution of claims, where it indicates that approximately 350,000 claims are labeled as “Not Fraud,” while more than 200,000 claims are classified as “Fraud”.

**Fig. 1.**
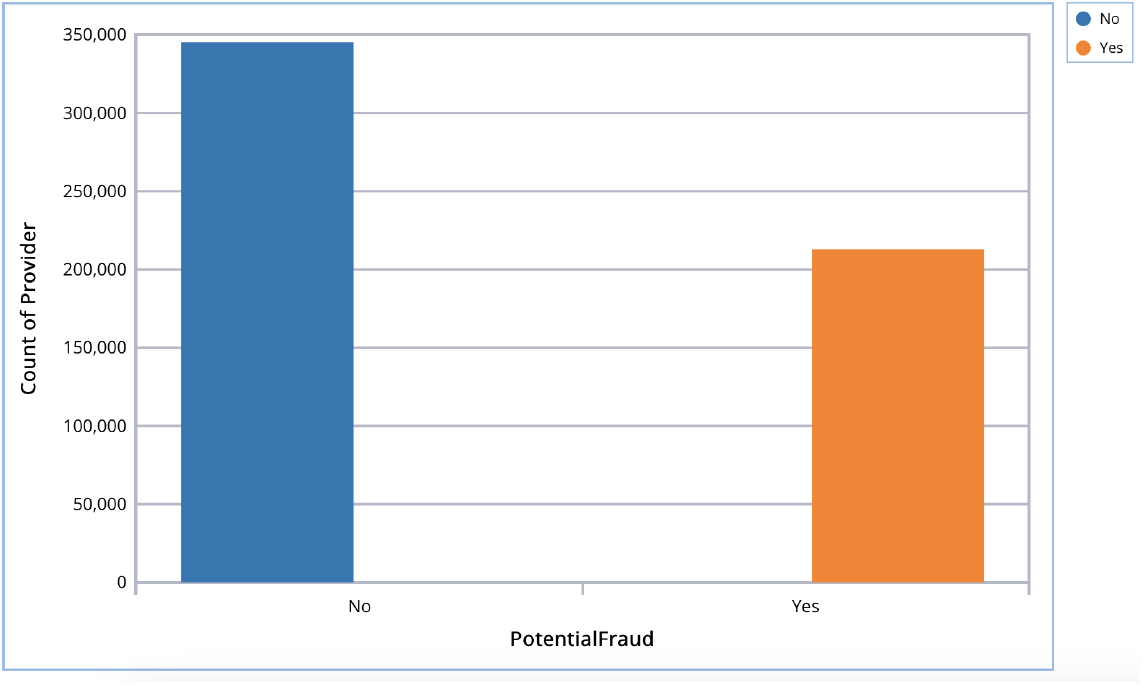
The claims distribution are categorized as “Potential Fraud” with two labels: “Yes” and “No”.

### 3.2 Feature Selection

Feature selection is the process of choosing a subset of features and analyzing only those features. This process becomes particularly important when dealing with high-dimensional datasets like Medicare. By choosing and retaining only the most relevant features, feature selection reduces the inclusion of less important ones during model training. This not only saves computational resources, but also improves the model ‘s performance, particularly in terms of accuracy. The dataset utilized in this research contains 56 features where all the features do not seem very relevant.

Therefore, during the experiment, the feature selection methods were employed to identify and choose the most relevant attributes. To do so, two filter-based feature selection methods were explored. One method is the Chi-Squared, and the other one is the Mutual Info [15]. Those two methods are well-known for categorical output variables based on the input variables, as depicted in Figure [2]. In the following sub-sections, the two feature selection methods, and the mathematics behind the process of choosing the best features for the proposed model will be elaborately discussed.

**Fig. 2.**
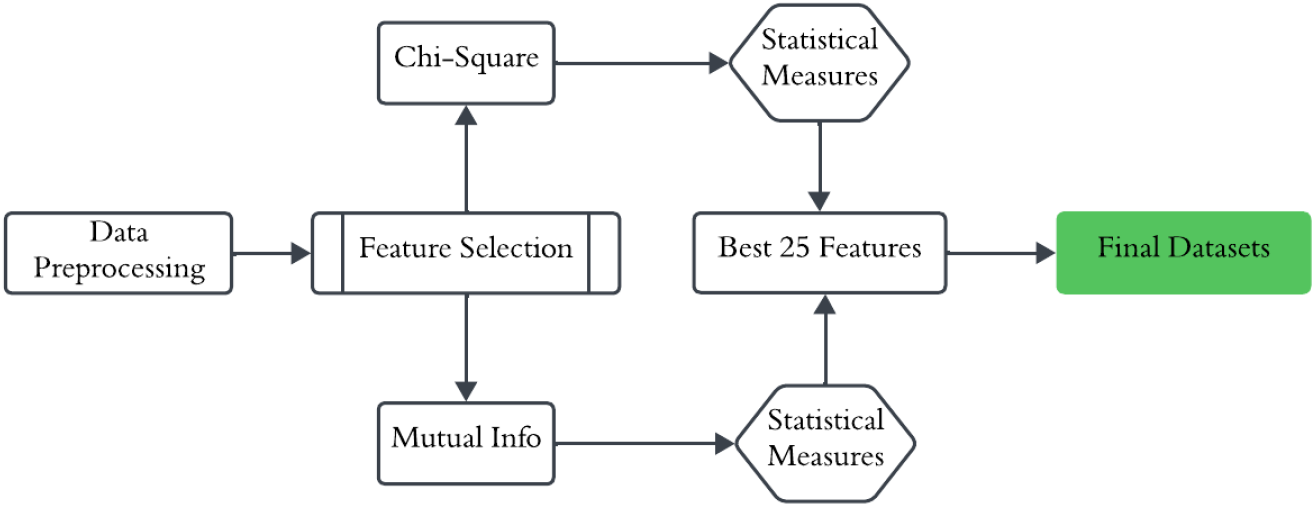
The feature selection process to select the best features based on a statistical measure.

#### 3.2.1 Chi-Squared Feature Selection

Chi-squared feature selection is a methodology that calculates the independence of each feature or attribute from the output class. This method is used in classification tasks with categorical output, such as a Binary classification. By comparing the observed and expected class frequencies, it determines the chi-squared statistic for each attribute, indicating the strength of the relationship. Equation (1)[16] describes how chi-squared values are calculated based on chi-squared statistics for each feature. After calculating the chi-squared statistics, it calculates the P-value, which characterizes the probability of observing a chi-square statistic. For each feature, having a lower P-value means a stronger connection with the target variable. In this experiment, the best 25 features based on the chi-squared feature importance were selected, as shown in Figure [3]. The experiment was implemented by the Scikit learn feature selection package by utilizing SelectKBest and chi2. Accordingly, the two variables with the highest Chi Square significance scores, provider_InscClaim_AmtReimbursedstd and provider_InscClaim_AmtReimbursedmean, indicate a strong correlation with the target variable, whether it is Fraud or Not Fraud.

**Fig. 3.**
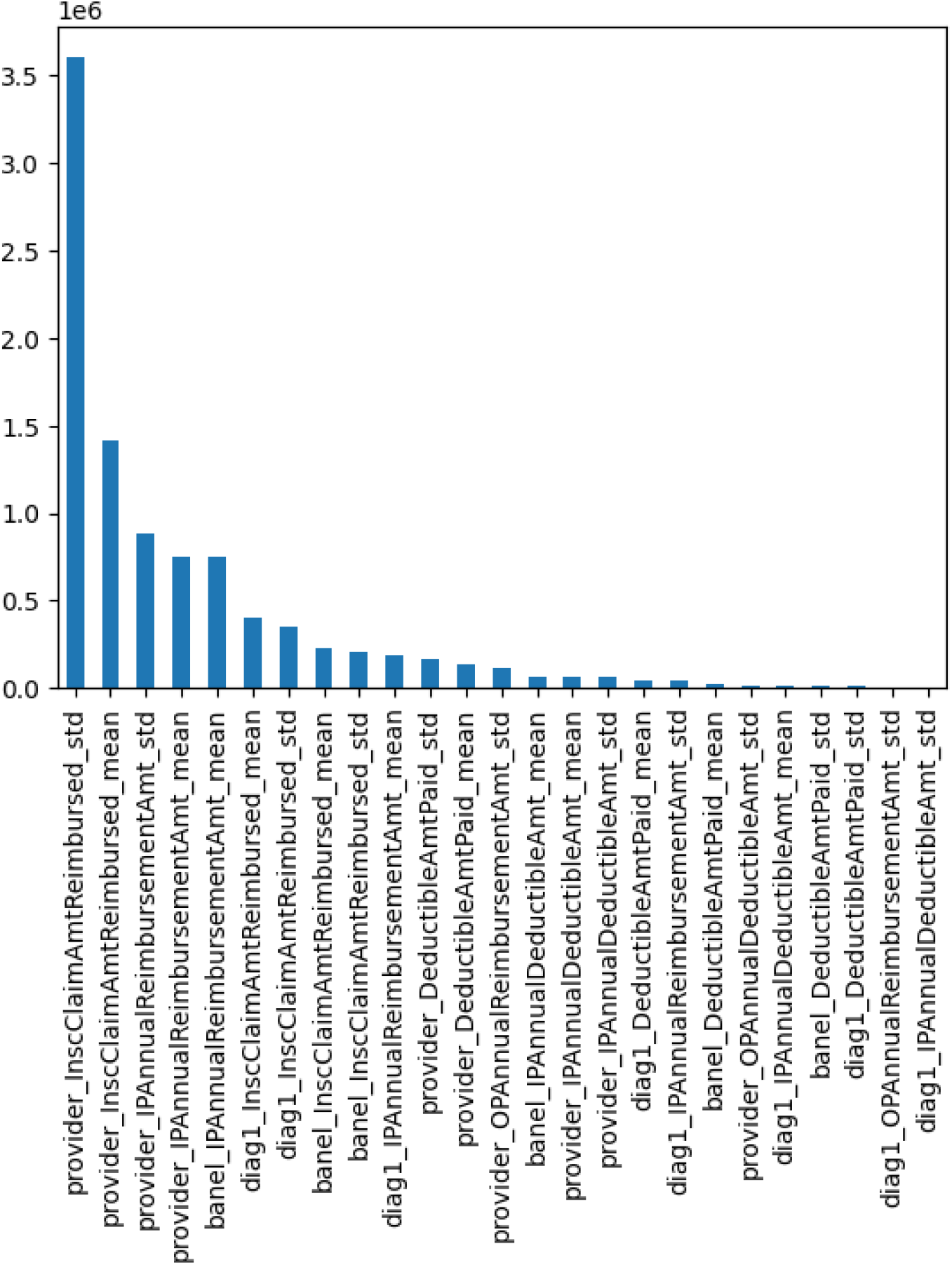
Top 25 features by using chi-squared feature selection methods.

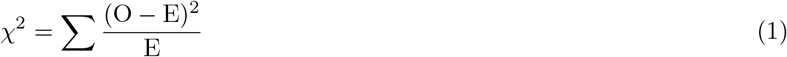

where,

*χ*^2^ is the chi-squared statistics.

O represents the observed frequency for each class in a feature.

E is the expected frequency.

#### 3.2.2 Mutual Info Feature Selection

Mutual info is a feature selection method that calculates the statistical dependence between a feature and the target variable. It describes how much information a feature contains about the target variable. For each feature, having a higher Mutual information score means a stronger connection with the target variable. The mutual information between a target variable (Y) and a feature (X) is calculated mathematically, as shown in the equation (2)[16].

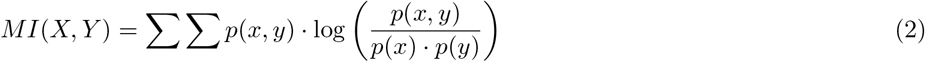

*Where*,

*MI*(*X, Y*) denotes the Mutual information between feature *X* and target *Y*.

*p*(*x, y*) illustrates the joint probability distribution of *X, Y*.

*p*(*x*) and *p*(*y*) are the marginal probability distributions.

To implement the Mutual info method in this research experiment, the Scikit learn feature selection package was used by utilizing SelectKBest and mutual info classif. The best 25 features were extracted from this experiment, which were used for training the model and recording observations regarding model accuracy. In this case, all the features got the same importance score of 0.024503, which demonstrates each feature has equal importance for predicting the target variable.

### 3.3 Data Sampling

Data sampling serves as a valuable approach for addressing class imbalance, effectively modifying the class distribution that encompasses both the majority and minority classes. This technique proves particularly advantageous when confronted with datasets where one class significantly outweighs the other. For the Medicare dataset, the majority class No (Not Fraud) constitutes 61.6% of the samples, while the minority class Yes (Fraud) comprises 38.4% of the samples (see Figure [4]). Different data sampling methods were employed in order to offset the class ratio for both of the classes. These methods include Random Under Sampling (RUS), Random Over Sampling (ROS), and Synthetic Minority Over Sampling technique (SMOTE)[17]. Different data sampling technique has different outcomes depending on the class ratio or classifier.

**Fig. 4.**
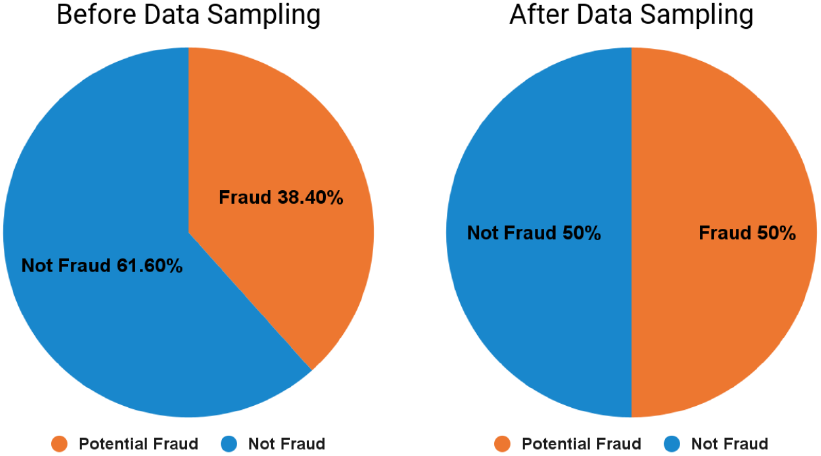
Data sampling technique to manipulate class imbalance.

The three mentioned data sampling techniques were examined and implemented in the Deep Learning model, which is the key focus of this paper.

#### 3.3.1 Random Under sampling (RUS)

Random under-sampling (RUS) is a technique that samples the data by randomly removing instances from the majority class. Many studies have shown that RUS performs very well compared to no data sampling [18]. As the Medicare dataset contains more samples from the “Not Fraudulent” class instead of “Potential Fraudulent”, the RUS was implemented using the imbalanced-learn library in Python to remove random numbers of samples from the “Not Fraudulent” class. The sampling strategy parameter was utilized to regulate the amount of data that needs to be removed. In the context of this research experiment, approximately 15% of the data originating from the majority class was deliberately omitted, thereby achieving a balanced 50:50 class distribution.

#### 3.3.2 Random Over sampling (ROS)

Another Data sampling technique that deployed in this study is Random Oversampling (ROS), which randomly adds instances to the minority class to balance the class ratio. Random Oversampling can lead to the duplication of sample data, creating instances where certain rows contain identical feature values. In the case of the Medicare dataset, which comprises thousands of data samples, applying random oversampling can generate additional rows with randomized data. The ROS was implemented using the imbalanced-learn library, which was applied for RUS, to balance the class ratio between Fraud and Not Fraud.

#### 3.3.3 Synthetic Minority Oversampling Technique (SMOTE)

The Synthetic Minority Oversampling technique (SMOTE) is a type of oversampling that creates brand-new artificial instances between minority instances that are close to one another. SMOTE operates by initially choosing a data point from the minority class, and then identifying its nearest neighbors within the same class. From these neighbors, one is randomly selected. Synthetic examples are subsequently generated by mixing the attributes of the chosen data point and neighbor with random values that stay within the original attribute ranges. This sequence is repeated to produce the necessary synthetic examples, thereby mitigating class distribution imbalance. Unlike ROS, this approach does not lead to data duplication. Consequently, this method can result in a more precise model with reduced overfitting.

### 3.4 Deep Learning and Proposed Model

Several research papers have concentrated mostly on traditional machine-learning techniques like random forest, logistic regression, or extreme gradient boosting to detect Medicare fraud. Nevertheless, this study shifts the focus to the Deep learning model to improve the model ‘s accuracy. A deep neural network model was developed using the Keras Sequential model with a few hidden layers. The input layer comes with a Rectified Linear Unit (Relu) activation function with a Dense layer where the output layer is connected with a sigmoid function. The sigmoid function maps the input values between a range of 0 and 1 to be used for binary classification problems. The intention is to keep the deep learning model straightforward. The neural network model was developed and trained using different feature selection and data sampling techniques while keeping the model parameters the same for all the experiments. Lastly, a combination of the best feature selection (Chi-square) and data sampling (SMOTE) method was proposed to train the model, as illustrated in Figure [5].

**Fig. 5.**
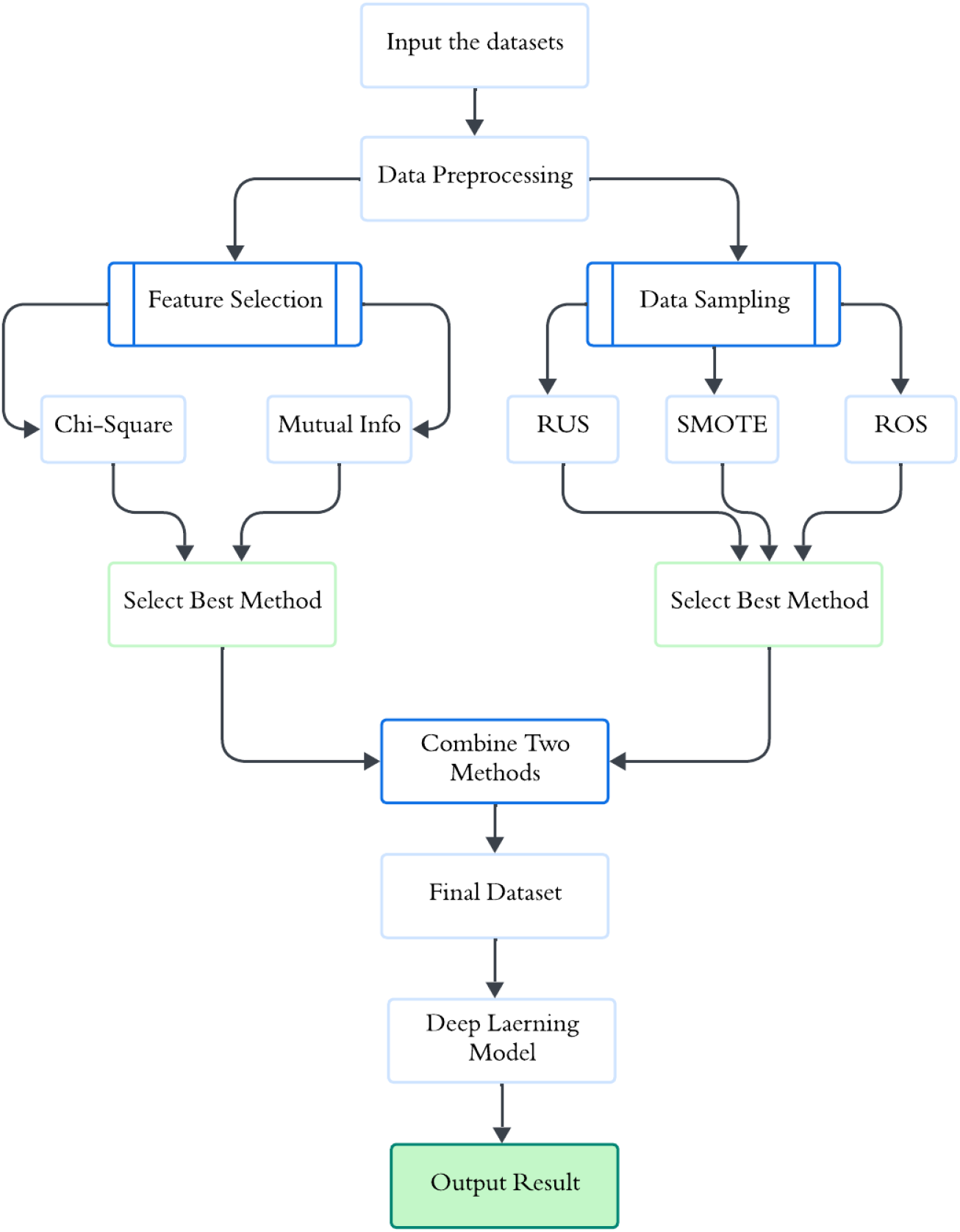
The process of improving the model performance by combining the feature selection and Data sampling techniques.

### 3.5 Code Availability

The custom code used for data preprocessing, feature selection, data sampling, and deep learning model training in this study is publicly available at: https://github.com/fahammed2022/Medicare_Fraud_Detection. To support reproducibility and long-term accessibility, a versioned snapshot of the code is archived in a DOI-minting repository (Zenodo): https://doi.org/10.5281/zenodo.18382713.

## 4 Results

In this section, a comprehensive analysis of the recorded performance findings derived from the proposed experiments is presented. Table 1 introduces the different accuracy percentages of distinct technique combinations, revealing the impact of applying feature selection and data sampling techniques.

**Table 1.** The result table indicates the accuracy of each experiment.

| Model | Feature Selection | Data Sampling | Accuracy |
| --- | --- | --- | --- |
| Baseline Model | NONE | NONE | 0.920 |
| Deep Learning Model | Chi Square (Best 25) | NONE | 0.903 |
| Deep Learning Model | Mutual Info (Best 25) | NONE | 0.895 |
| Deep Learning Model | NONE | RUS | 0.914 |
| Deep Learning Model | NONE | ROS | 0.943 |
| Deep Learning Model | NONE | SMOTE | 0.957 |
| Proposed Model | Chi Square (Best 25) | SMOTE | 0.954 |

To begin with, a baseline deep learning model was experimented without employing any of the previously mentioned techniques (feature selection or data sampling), resulting in an accuracy of 0.920. Using the feature selection technique, the accuracy of selecting the top 25 features applying Chi-square recorded an accuracy of 0.903. At the same time, it experienced a slight decrease, dropping to 0.895 by using Mutual Info.

On the other hand, the accuracy increased drastically when different data sampling methods were employed, especially ROS and SMOTE. By implementing RUS, the recorded accuracy is 0.914, which is slightly lower than the baseline model. In contrast, 0.943 accuracy is obtained by using the ROS. The most significant improvement is witnessed with the SMOTE, which yields an accuracy of 0.957.

In the final comparison stage, the proposed scheme was evaluated by combining the best techniques in terms of the highest accuracy rate obtained, which are Feature selection (Chi-squared) and data sampling (SMOTE). The attained accuracy result is 95.4%, remarkably similar to SMOTE with 95.7%. The suggested deep learning model, along with the combined techniques, performed exceptionally well for both the training and validation sets of data, as shown in the learning curve Figure [6]. The training accuracy was nearly 98%, while the validation obtained an accuracy of around 95.5%.

**Fig. 6.**
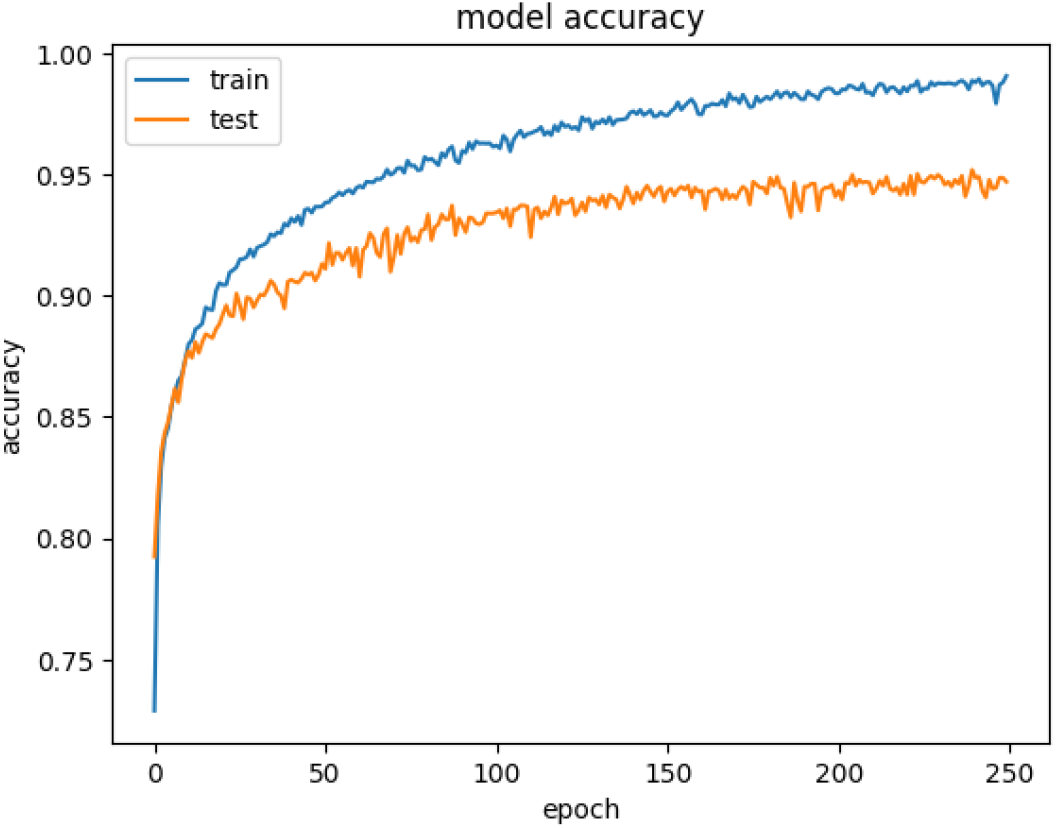
The learning curve of the proposed model.

To provide a more comprehensive evaluation of the proposed model ‘s efficiency, a detailed analysis of performance metrics is presented in Table 2. The results indicate that the combination of Chi-squared feature selection and SMOTE sampling achieved a high F1-score of 0.94 for both classes, demonstrating balanced performance despite the inherent class imbalance in Medicare data. Specifically, the model reached a Recall of 0.98 for the ‘Fraud’ class, correctly identifying 98% of all fraudulent instances. This indicates that the proposed model effectively minimizes False Negatives, which is a critical priority in healthcare fraud detection to protect financial resources and maintain system integrity. Furthermore, the consistency between the 95.4% Accuracy and the 94% F1-score confirms the stability and reliability of the integrated deep learning framework.

**Table 2.**
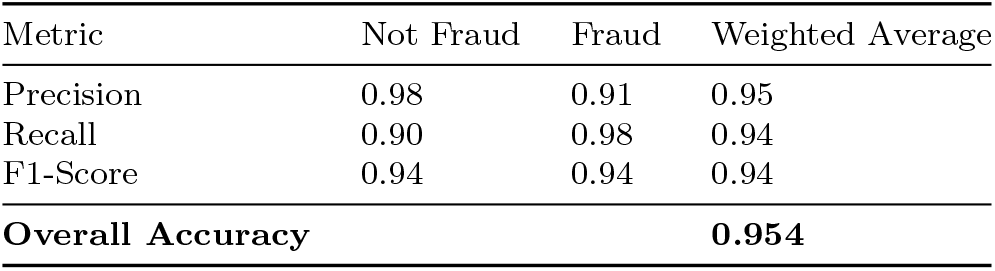
Detailed performance metrics for the Proposed Model (Chi-Square + SMOTE).

Accordingly, when considering the choice of the most suitable model regarding Medicare fraud detection, the obtained results tend to favor the combined model. This selection is based on its demonstrated attributes of reduced overfitting and consistently stable accuracy compared to the other models explored in this research.

Finally, The minimal gap between the training and validation accuracy shown in the learning curve Figure [6] serves as a proxy for the model ‘s reliability, indicating low variance and consistent performance across different data partitions. This stability suggests that the reported accuracy values are robust and less prone to significant variability when applied to similar Medicare claim distributions.

## 5 Conclusion

The Medicare fraud phenomenon presents an ever-evolving challenge that signals urgency for a sophisticated detection model. The existing solutions rely on conventional approaches; however, the proposed model in this study deviates from this norm by leveraging the capability of a deep learning model along with data sampling and feature selection techniques. The primary aim of this integration is to improve the model ‘s accuracy as well as performance notably. The observed result from the experiment depicts that the integration between Chi-square and SMOTE techniques indicates an outstanding ability to detect fraudulent claims with 95.4% accuracy, with insignificant evidence of overfitting detected. This finding advocates the shift to manipulate integrated techniques instead of just employing a baseline deep learning model for better results when detecting Medicare fraud. This paper sparks a set of recommendations for future work, such as experimenting with the proposed model on a diverse Medicare dataset to refine and validate the model ‘s efficacy. Also, conducting further experiments to include a diverse array of feature selection and data sampling techniques along with specific data sampling ratios (e.g., 65:30 and 75:25). Lastly, integrating the proposed model with an emerging technology, such as the blockchain will be a game-changer in the way the medical record is managed and stored. The blockchain technology is practically feasible if the decentralized ledger is used as the primary data generation layer. In this setup, the blockchain ensures that medical records are tamper-proof before they even reach the deep learning model. This approach creates a clear audit trail and ensures the deep learning model works with verified data, which significantly improves the security of the fraud detection process.

## Data Availability

All data produced in the present study are available upon reasonable request to the authors

https://www.kaggle.com/datasets/rohitrox/healthcare-provider-fraud-detection-analysis

## Declarations

### Limitation

While this study focuses on the U.S. Medicare dataset, the framework requires validation against international datasets to confirm its efficiency across global insurance systems. The reliance on historical data also introduces a temporal limitation, as fraudulent patterns are constantly evolving. Furthermore, while the current architecture is highly reliable, the computational resources required for real-time detection on larger, live-streaming datasets remain a challenge that requires further optimization. Finally, a significant constraint for broader use is the dependency on granular, provider-level feature engineering, which may vary in environments with different reporting standards.

### Ethical Approval

Not applicable.

### Availability of data and materials

The dataset used for this experiment is publicly available on kaggle.com. Source: https://www.kaggle.com/datasets/rohitrox/healthcare-provider-fraud-detection-analysis.

### Authors’ contributions

FA: Contributed significantly to the idea and design of the work, collected and prepared the dataset for the experiments, carried out the experiments, gave technical guidance and support, wrote some of the sections of the manuscript such as methodologies and results, and extensively revised the completed work and the final manuscript. BAB: made a major contribution to the idea and design of the work, carried out a review of the related work, gave technical guidance and support, helped write up the rest of the manuscript, and extensively revised the completed work and the final manuscript. OM: Made a substantial contribution to the concept and structure of the work, suggested the relevant literature, and carefully reviewed the final manuscript and all of its drafts for meaningful academic value.

## References

[1] U.S. Department of Justice: 2021 National Health Care Fraud Enforcement Action. Accessed: 2023-10-25 (2021). https://www.justice.gov/criminal-fraud/2021-nhcf/district-summaries

[2] Li, J., Lan, Q., Zhu, E., Xu, Y., Zhu, D.: A study of health insurance fraud in china and recommendations for fraud detection and prevention. Journal of Organizational and End User Computing (JOEUC) 34(4), 1–19 (2022)

[3] Xu, B., Wang, Y., Liao, X., Wang, K.: Efficient fraud detection using deep boosting decision trees. Decision Support Systems, 114037 (2023)

[4] Kapadiya, K., Patel, U., Gupta, R., Alshehri, M.D., Tanwar, S., Sharma, G., Bokoro, P.N.: Blockchain and ai-empowered healthcare insurance fraud detection: An analysis, architecture, and future prospects. IEEE Access 10, 79606–79627 (2022)

[5] Vyas, S., Serasiya, S., Vyas, A.: Combined approach of ml and blockchain for fraudulent detection in insurance claim. In: 2022 International Conference on Edge Computing and Applications (ICECAA), pp. 544–550 (2022). IEEE

[6] Chen, J.-P., Lu, P., Yang, F., Chen, R., Lin, K.: Medical insurance fraud detection using graph neural networks with spatio-temporal constraints. Journal of Network Intelligence 7, 480–498 (2022)

[7] Zhang, G., Zhang, X., Bilal, M., Dou, W., Xu, X., Rodrigues, J.J.: Identifying fraud in medical insurance based on blockchain and deep learning. Future Generation Computer Systems 130, 140–154 (2022)

[8] Akbar, N.A., Sunyoto, A., Arief, M.R., Caesarendra, W.: Improvement of decision tree classifier accuracy for healthcare insurance fraud prediction by using extreme gradient boosting algorithm. In: 2020 International Conference on Informatics, Multimedia, Cyber and Information System (ICIMCIS), pp. 110–114 (2020). IEEE

[9] Settipalli, L., Gangadharan, G.: Wmtdbc: An unsupervised multivariate analysis model for fraud detection in health insurance claims. Expert Systems with Applications 215, 119259 (2023)

[10] Ataabadi, P.E., Neysiani, B.S., Nogorani, M.Z., Mehraby, N.: Semi-supervised medical insurance fraud detection by predicting indirect reductions rate using machine learning generalization capability. In: 2022 8th International Conference on Web Research (ICWR), pp. 176–182 (2022). IEEE

[11] Herland, M., Bauder, R.A., Khoshgoftaar, T.M.: The effects of class rarity on the evaluation of supervised healthcare fraud detection models. Journal of Big Data 6, 1–33 (2019)

[12] Massi, M.C., Ieva, F., Lettieri, E.: Data mining application to healthcare fraud detection: a two-step unsupervised clustering method for outlier detection with administrative databases. BMC medical informatics and decision making 20, 1–11 (2020)

[13] Parnian, K., Sorouri, F., Souha, A.N., Molazadeh, A., Mahdavi, S.: Fraud detection in health insurance using a combination of feature subset selection based on squirrel optimization algorithm and nearest neighbors algorithm methods. Future Gener Distrib Syst J 3(2), 1–11 (2021)

[14] Gupta, R.A.: Healthcare Provider Fraud Detection Analysis. https://www.kaggle.com/datasets/rohitrox/healthcare-provider-fraud-detection-analysis. Accessed: 2023-09-22 (2018)

[15] Li, J., Cheng, K., Wang, S., Morstatter, F., Trevino, R.P., Tang, J., Liu, H.: Feature selection: A data perspective. ACM computing surveys (CSUR) 50(6), 1–45 (2017)

[16] Khah, S.S., Wu, Y.: An enhanced ad event-prediction method based on feature engineering. arXiv preprint arXiv:1907.01959 (2019)

[17] Rendon, E., Alejo, R., Castorena, C., Isidro-Ortega, F.J., Granda-Gutierrez, E.E.: Data sampling methods to deal with the big data multi-class imbalance problem. Applied Sciences 10(4), 1276 (2020)

[18] Seiffert, C., Khoshgoftaar, T.M., Van Hulse, J., Folleco, A.: An empirical study of the classification performance of learners on imbalanced and noisy software quality data. Information Sciences 259, 571–595 (2014)

